# The impact of patient-centered care on quality of life and hope among patients receiving home medical care: The Zaitaku Evaluative Initiatives and Outcome Study

**DOI:** 10.1101/2023.12.07.23299634

**Authors:** Hidekazu Iida, Shinu Hayashi, Masakazu Yasunaka, Yukio Tsugihashi, Misaki Hirose, Yutaka Shirahige, Noriaki Kurita, the ZEVIOUS group

## Abstract

**Objectives:** This study aimed to examine the correlation between the quality of patient-centered care and quality of life and hope among patients receiving home medical care.

**Design:** Multicenter cross-sectional study

**Setting and Participants:** This study was part of the Zaitaku Evaluative Initiatives and Outcome Study involving 29 home care clinics in Japan. The participants were patients receiving home medical care who were judged capable of responding to the questionnaire survey by their attending physician.

**Methods:** Patient centeredness, the exposure variable, was measured using the Japanese version of the Primary Care Assessment Tool–Short Form (JPCAT-SF). Outcomes, namely quality of life and hope, were measured utilizing the Quality of Life-Home Care (QOL-HC) and Health-Related Hope (HR-Hope) scales, respectively. Mixed-effects linear regression models were applied, incorporating covariates such as age, sex, education, family member presence, comorbidities (primary and other), depressive symptoms, residence type, and patient life expectancy.

**Results:** Among the 194 participants, a notable association was found, wherein a higher JPCAT-SF total score correlated with an elevated QOL-HC score (adjusted mean difference per 10-point increase: 0.29, 95% CI: 0.17–0.42). Within the JPCAT-SF domains, elevated scores for first contact, longitudinality, comprehensiveness, and community orientation were correlated with higher QOL-HC scores. Additionally, a higher JPCAT-SF total score was associated with elevated HR-Hope levels (adjusted mean difference per 10-point increase: 5.1, 95% CI: 3.2–7). Higher scores for first contact, longitudinality, coordination, comprehensiveness, and community orientation were associated with higher HR-Hope scores.

**Conclusions and Implications:** The findings underscore that higher quality patient-centered care is positively associated with enhanced quality of life and hope among home medical care patients. This study highlights the importance of strengthening patient centeredness in daily clinical practice.

## Introduction

Maintaining or improving function and well-being are central goals in caring for patients requiring home medical care when patients have chronic or progressive illnesses for which there is no cure or for which treatment is not feasible.^1^ Thus, patient-reported outcomes (PROs), which reflect quality of life (QOL) and hope, are essential targets for in-home medical care. Nevertheless, QOL among patients receiving home medical care is often underestimated owing to such patients’ limited activities of daily living (ADLs).^2^ Empirical evidence on approaches that home medicine physicians can implement to improve their patients’ QOL is scarce compared with approaches for outpatient and nursing home settings. For example, patient-centered care for patients with type 2 diabetes was associated with physical and mental health related-QOL.^3^ In nursing home settings, patient-centered care, such as building close relationships and collaborative decision-making, was associated with better QOL.^4^

Hope is considered an essential coping strategy,^5^ given that it is the only thing that patients can do in a clinical oncology setting when they may seem to be unable to do anything.^6^ The benefit of hope is supported by empirical evidence showing that higher hope is associated with lesser pain and psychological distress among patients with lung cancer.^7^ Furthermore, hope has been identified as an essential theme of patient-centered care for chronic illnesses.^8^ Despite the importance of QOL and the potential of hope in improving patient outcomes, there is a shortage of empirical research showing that providing patient-centered care improves these outcomes among patients receiving home medical care.^8^

In-home medical care for adults with mental health problems in the United Kingdom, providing elements constituting patient-centered care, such as sufficient time to talk and resolving patients’ problems, was positively correlated with hope.^9^ However, patient-centered care comprises multiple broad domains such as first contact (refers to access to medical care, including availability outside regular hours or on days off) and longitudinality (refers to understanding the whole person and not just the disease).^10^ In addition, whether patient-centered care can ensure the maintenance of QOL and hope among patients with impaired physical functioning due to chronic illness rather than psychological problems has not been examined. Clarifying this issue could contribute to further empirical work leading to an understanding of how attending physicians can provide patient-centered home medical care.^8^

Therefore, we conducted a multicenter cross-sectional study using data from the Zaitaku Evaluative Initiatives and Outcome Study (ZEVIOUS) to examine the association of the quality of patient-centered care with QOL and hope among patients receiving home medical care.

## Methods

### Design, Setting, and Participants

This study was part of ZEVIOUS, a multicenter cross-sectional study involving 29 home care clinics in the Tokyo Metropolitan area, Nara Prefecture, and Nagasaki Prefecture in Japan. The inclusion criteria were patients receiving home medical care, as determined by their attending physicians, and judged capable of responding to the questionnaire survey. The questionnaire was distributed to each patient between January and July 2020, and the patients were requested to complete the survey. Patients with visual impairments or physical disabilities that prevented them from writing were permitted assistance by a family member or formal caregiver in answering the survey. The study protocol was approved by the Institutional Review Board of Fukushima Medical University.

### Patient centeredness as an Exposure

The Japanese version of the Primary Care Assessment Tool–Short Form (JPCAT-SF) was used to measure patient centeredness in primary care settings.^10^ The JPCAT-SF comprises 13 items encompassing six domains representing five primary care attributes: first contact (two items), longitudinality (two items), coordination (three items), comprehensiveness (two items for “services available” and two items for “services provided”), and community orientation (two items). Detailed information regarding the items, scoring, psychometric properties, and domains of the JPCAT-SF can be found in the supplementary information (Table S1, Text S1). Participants rated each item on a five-point Likert scale ranging from strongly disagree to strongly agree. We converted the responses to item scores ranging from 0 to 4 points. Domain scores were calculated by multiplying the average item scores within the same domain by 25, resulting in a range of 0–100 points, with higher scores indicating better performance. The total score represents an overall measure of the patient centeredness of primary care and was calculated as the average of the six domain scores.

### QOL and Hope as Outcomes

The Quality of Life-Home Care (QOL-HC) is a four-item questionnaire that assesses the QOL of older patients receiving home medical care.^11^ Kamitani et al. demonstrated the face validity of the QOL-HC through item derivation by physicians and care managers and item selection by geriatricians.^11^ Each item is rated on a three-point scale, ranging from “never agree” (0 points) to “always agree” (2 points), resulting in a total score ranging from 0 to 8 points. (Table S2)

The Health-Related Hope (HR-Hope) Scale is an 18-item, unidimensional scale designed to evaluate HR-Hope among individuals with chronic conditions.^12^ Through structural validation, the scale comprises three subdomains: “something to live for” (five items), “health and illness” (six items), and “role and connectedness” (seven items) (Supplementary Table).^12^ Participants rate their responses to each item on a four-point Likert scale ranging from 1 = I do not feel that way at all to 4 = I strongly feel that way. After obtaining the average score for each subdomain and the total score, the scores are scaled from 0 to 100 points. Patients without family were exempted from answering two items in the “role and connectedness” subdomain.(Table S3)

### Covariates

Demographic information, such as age, sex, education, and the presence of family members, was collected through a questionnaire. The physician-in-charge provided data on comorbidities, type of residence, and patient life expectancy. Regarding the patient’s life expectancy, the home medical care physician assigned to the patient answered the following question: “What diseases were the leading cause of introducing home medical care?” The physicians were allowed to choose from the following options: cerebrovascular disease, heart disease, cancer, respiratory disease, joint disease, dementia, incurable neuromuscular disease, diabetes, visual and hearing impairment, fractures and falls, spinal cord disease, infirmity, other, and unknown. Regarding type of residence, the physicians answered the question: “What is the type of residence?” The following options were provided: home, care home for older adults, retirement home, group home (for patients with dementia), multifunctional long-term care in a small group home, and short stay. We classified the responses into those with homes and those without homes (nursing homes). Lastly, physicians were asked “How long do you expect the clinical prognosis (life expectancy) of this patient to be?” They were allowed to choose from five options: “less than one month,” “more than one month to less than three months,” “more than three months to less than six months,” “more than six months to less than 12 months,” and “more than 12 months.” Depressive symptoms were assessed using the five-question Mental Health Inventory (MHI-5).^13^ The MHI-5 scores were calculated according to a previous study, and the total score was converted to 0–100. A score of ≤52 on the MHI-5 was defined as having depression.^13^

### Statistical Analysis

Statistical analyses were performed using Stata/SE version 15 (StataCorp, College Station, TX, USA). Patient characteristics were described using means and standard deviations for continuous variables and frequencies and percentages for categorical variables. Mixed-effects linear regression models were utilized to estimate the association between JPCAT-SF scores and QOL-HC and HR-Hope scores, considering clustering effects by the facility. Robust variance estimation was used for the QOL-HC analysis because the scale did not meet the standard assumptions of equal variance and normality. Age, sex, educational attainment, family presence, depressive symptoms, patient life expectancy, and comorbidities were included as covariates in the models.

Additionally, the models were fitted, in which each of the six domain scores of the JPCAT-SF was treated as an explanatory variable. Missing covariate data were addressed using multiple imputations with chained equations, assuming that the mechanism of missing data was random. The imputed data were analyzed five times.^14^ Statistical significance was defined as p < 0.05.

## Result

### Patient Characteristics

Of the 202 patients who received home medical care, eight without a JPCAT-SF score were excluded, resulting in a final sample size of 194 patients for analysis. Patient characteristics are presented in Table 1.

**Table 1.**
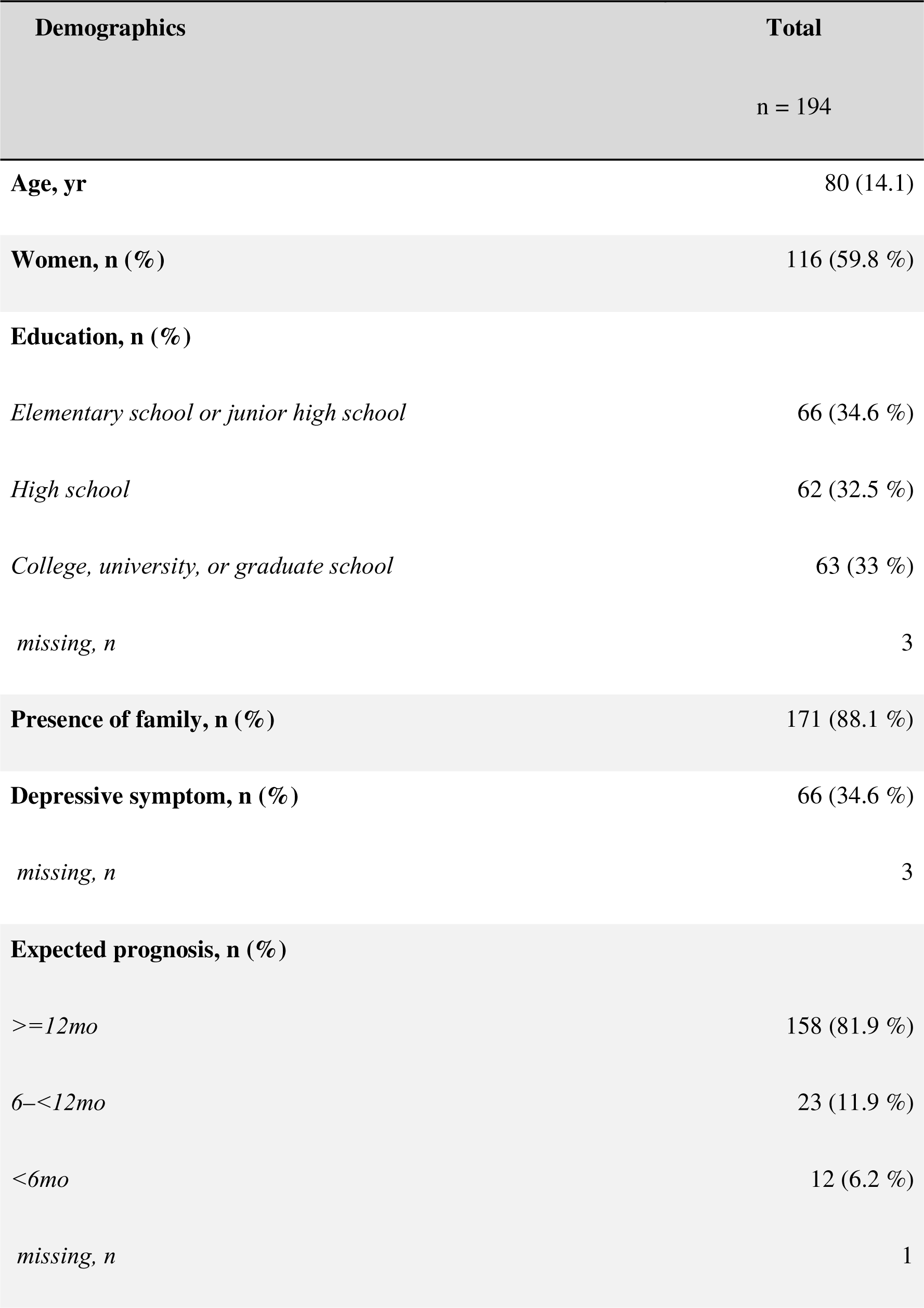
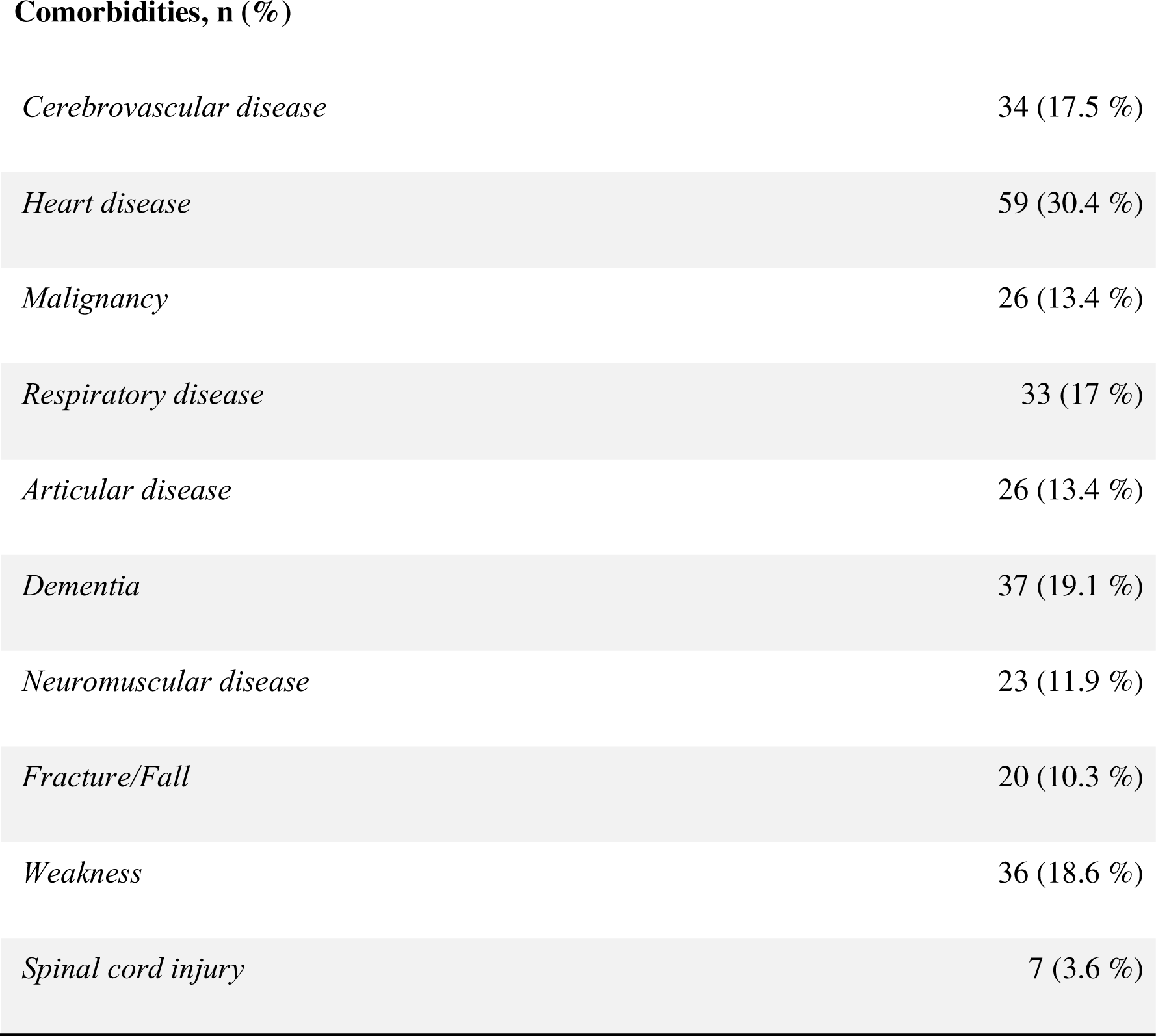
Characteristics of patients (n= 194)

| **Demographics** | **Total** |
| --- | --- |
|  | n = 194 |
| **Age, yr** | 80 (14.1) |
| **Women, n (%)** | 116 (59.8 %) |
| **Education, n (%)** |  |
| *Elementary school or junior high school* | 66 (34.6 %) |
| *High school* | 62 (32.5 %) |
| *College, university, or graduate school* | 63 (33 %) |
| *missing, n* | 3 |
| **Presence of family, n (%)** | 171 (88.1 %) |
| **Depressive symptom, n (%)** | 66 (34.6 %) |
| *missing, n* | 3 |
| **Expected prognosis, n (%)** |  |
| *>=12mo* | 158 (81.9 %) |
| *6-<12mo* | 23 (11.9 %) |
| *<6mo* | 12 (6.2 %) |
| *missing, n* | 1 |
| **Comorbidities, n (%)** |  |
| *Cerebrovascular disease* | 34 (17.5 %) |
| *Heart disease* | 59 (30.4 %) |
| *Malignancy* | 26 (13.4 %) |
| *Respiratory disease* | 33 (17 %) |
| *Articular disease* | 26 (13.4 %) |
| *Dementia* | 37 (19.1 %) |
| *Neuromuscular disease* | 23 (11.9 %) |
| *Fracture/Fall* | 20 (10.3 %) |
| *Weakness* | 36 (18.6 %) |
| *Spinal cord injury* | 7 (3.6 %) |

### Association between JPCAT-SF, QOL-HC, and HR-Hope

Table 2 presents the association between the JPCAT-SF total score and QOL-HC. Higher total JPCAT-SF scores were associated with higher QOL-HC scores (adjusted mean difference for every 10-point increase: 0.29, 95% CI: 0.17–0.42). Age was also positively associated with QOL-HC (every ten years increase: 0.18, 95% CI: 0.05–0.3). Depressive symptoms were negatively associated with QOL-HC (−0.89, 95% CI: −1.36–-0.43). Sex, education level, family presence, expected prognosis, and comorbidities were not significantly associated with QOL-HC. The associations between each JPCAT-SF domain and QOL-HC are presented in Table 3. Among the JPCAT-SF domains, higher scores in first contact (0.15, 95% CI: 0.06–0.25), longitudinality (0.21, 95% CI: 0.10–0.31), comprehensiveness (services available: 0.11, 95% CI: 0.03–0.20; services provided: 0.08, 95% CI: 0.03–0.13), and community orientation (0.10, 95% CI: 0.02–0.18) were associated with higher QOL-HC scores, whereas coordination (0.03, 95% CI: −0.03–0.10) showed a non-significant association.

**Table 2.** Associations between patient experience and QOL-HC^a^ (n = 189)

| **QOL-HC, points** | **Mean difference (95% CI)** | **P-value** |
| --- | --- | --- |
| *JPCAT-SF, total, per 10 pt* | *0.29 (0.17 - 0.42)\** | *<0.001* |
| *Age, per 10 y* | *0.18 (0.05 - 0.3)\** | *0.005* |
| *Women vs. Men* | *0.18 (-0.24 - 0.6)* | *0.398* |
| *Educational attainment* |  |  |
| *Junior high school or lower* | *-0.17 (-0.64 - 0.3)* | *0.474* |
| *High school* | *-0.31 (-0.69 - 0.07)* | *0.114* |
| *College/University/Graduate school/Other* | *Reference* |  |
| *Presence of family* | *0.01 (-0.53 - 0.55)* | *0.974* |
| *Expected prognosis* |  |  |
| *>=12 month* | *Reference* |  |
| *6-<12 month* | *0.26 (-0.15 - 0.67)* | *0.214* |
| *<6 month* | *0.43 (-0.15 - 1.01)* | *0.145* |
| *Depressive symptom* | *-0.89 (-1.36 - -0.43)\** | *<0.001* |
| *Comorbidities* |  |  |
| *Cerebrovascular disease* | *0.12 (-0.57 - 0.8)* | *0.742* |
| *Heart disease* | -0.06 (-0.41 - 0.29) | 0.725 |
| *Malignancy* | -0.38 (-0.8 - 0.04) | 0.074 |
| *Respiratory disease* | 0.44 (-0.08 - 0.97) | 0.100 |
| *Articular disease* | -0.2 (-0.8 - 0.41) | 0.525 |
| *Dementia* | 0.38 (-0.11 - 0.86) | 0.128 |
| *Neuromuscular disease* | -0.09 (-0.66 - 0.49) | 0.768 |
| *Fracture/Fall* | -0.15 (-0.78 - 0.48) | 0.644 |
| *Weakness* | -0.26 (-0.78 - 0.25) | 0.315 |
| *Spinal cord injury* | 0.13 (-0.62 - 0.87) | 0.740 |

Analysis of 189 patients among 29 facilities.
<sup>a</sup>Mixed-effects linear regression model adjusted for covariates listed above.
**\*statistically significant**

**Table 3.** Associations between JPCAT-SF domain and QOL-HC^a^ (n = 189)

| QOL, points | Mean difference (95% CI) | P-value |
| --- | --- | --- |
| *First contact, per 10 pt* | *0.15 (0.06–0.25)\** | *0.0001* |
| *Longitudinality, per 10 pt* | *0.21 (0.10–0.31)\** | *0.00* |
| *Coordination, per 10 pt* | *0.03 (-0.03–0.10)\** | *0.30* |
| *Comprehensiveness (needs service), per 10 pt* | *0.11 (0.03–0.20)\** | *0.008* |
| *Comprehensiveness(service), per 10 pt* | *0.08 (0.03–0.13)\** | *0.002* |
| *Community Orientation, per 10 pt* | *0.10 (0.02–0.18)\** | *0.014* |

Analysis of 189 patients among 29 facilities.
<sup>a</sup>Mixed-effects linear regression model adjusted for covariates listed in Table 2.
**\*statistically significant**

Table 4 shows the association between the JPCAT-SF total score and HR-Hope. Higher total JPCAT-SF scores were associated with higher levels of HR-Hope (adjusted mean difference for every 10-point increase: 5.1, 95% CI: 3.2–7). Negative associations with HR-Hope were observed for expected prognosis (−16, 95% CI: −29–-3) and depressive symptoms (−12.9, 95% CI: −18.9–-6.9). Sex, education level, family presence, and comorbidities were not significantly associated. The associations between each JPCAT-SF domain and HR-Hope are presented in Table 5. Higher scores in first contact (2.6, 95% CI: 1.19–4.18), longitudinality (2.7, 95% CI: 1.07–4.43), coordination (1.2, 95% CI: 0.13–2.36), comprehensiveness (services available: 1.9, 95% CI: 0.55–3.38; services provided: 1.5, 95% CI: 0.07–2.38), and community orientation (2.4, 95% CI: 1.13–3.79) were associated with higher QOL-HC scores.

**Table 4.** Associations between JPCAT-SF and HR-Hope^a^ (n = 194)

| HR-Hope, points | Mean difference (95% CI) | P-value |
| --- | --- | --- |
| *JPCAT-SF, total, per 10 pt* | 5.1 (3.2 - 7)* | <0.001 |
| Age, per 10 y | -0.8 (-3.2 - 1.6) | 0.512 |
| Women vs. Men | 1.3 (-5 - 7.6) | 0.681 |
| Educational attainment |  |  |
| *Junior high school or lower* | -2.6 (-9.9 - 4.7) | 0.484 |
| *High school* | -6.6 (-14 - 0.7) | 0.077 |
| *College/University/Graduate school/Other* | Reference |  |
| Presence of family | -4.6 (-13.2 - 4.1) | 0.303 |
| Expected prognosis |  |  |
| *>=12 month* | Reference |  |
| *6-<12 month* | -3 (-12.5 - 6.5) | 0.532 |
| *<6 month* | -16 (-29 - -3)* | 0.016 |
| *Depressive symptom* | -12.9 (-18.9 - -6.9)* | <0.001 |
| Comorbidities |  |  |
| *Cerebrovascular disease* | -5.7 (-13.6 - 2.1) | 0.149 |
| *Heart disease* | 2.9 (-3.6 - 9.5) | 0.383 |
| *Malignancy* | 6.3 (-3.3 - 15.9) | 0.201 |
| *Respiratory disease* | -3.6 (-11.3 - 4.1) | 0.364 |
| *Articular disease* | -3.1 (-11.6 - 5.4) | 0.477 |
| *Dementia* | 4.3 (-3.6 - 12.3) | 0.285 |
| *Neuromuscular disease* | -1.4 (-11.3 - 8.5) | 0.786 |
| *Fracture/Fall* | 3.9 (-5.6 - 13.5) | 0.419 |
| *Weakness* | -1.6 (-10.3 - 7) | 0.714 |
| *Spinal cord injury* | -5.8 (-21.9 - 10.3) | 0.481 |

Analysis of 194 patients among 29 facilities.
Mixed-effects linear regression models adjusted for covariates listed above.
**\*statistically significant**

**Table 5.**
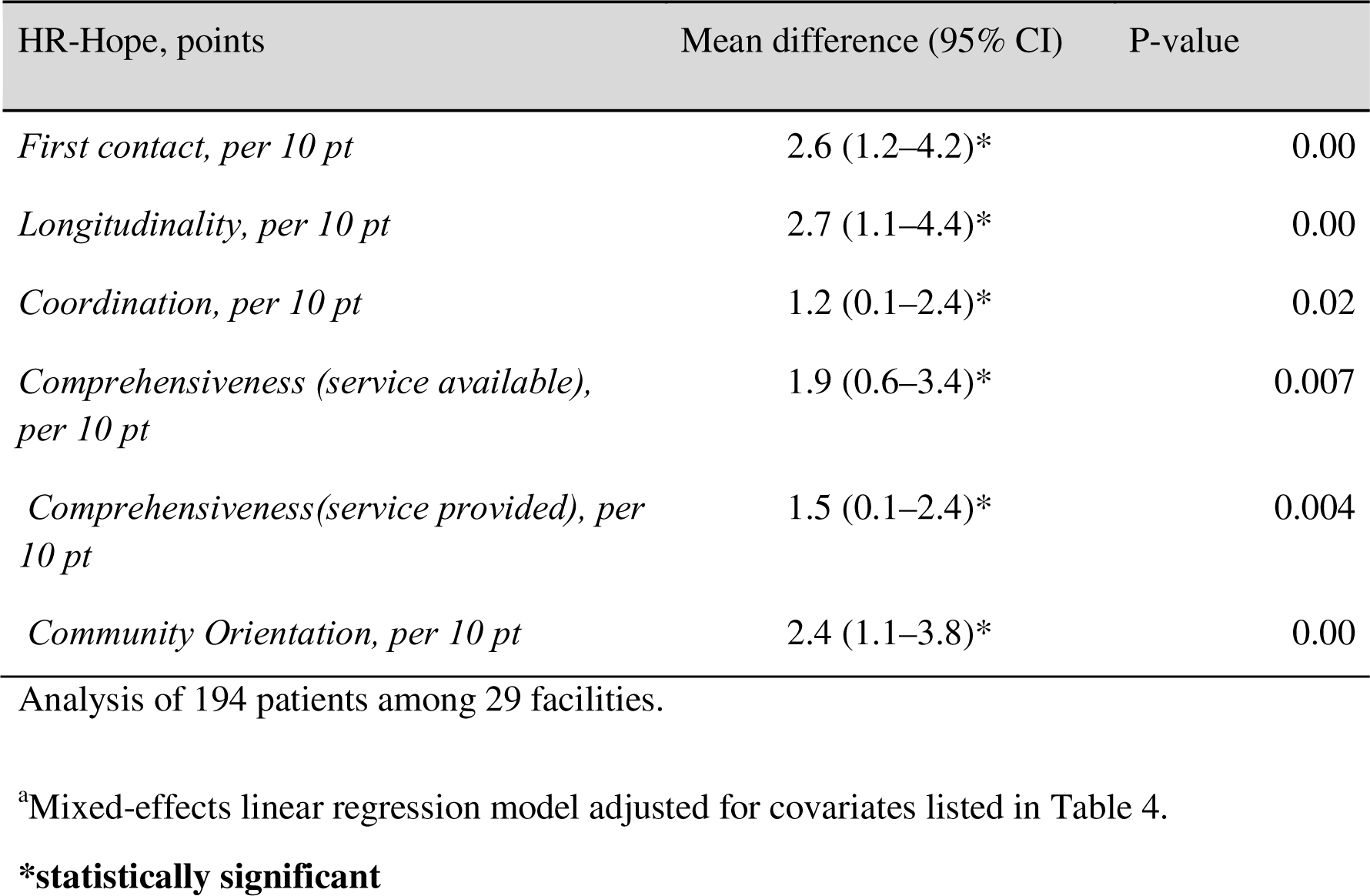
Associations between JPCAT-SF domain and HR-Hope^a^ (n = 194)

| HR-Hope, points | Mean difference (95% CI) | P-value |
| --- | --- | --- |
| *First contact, per 10 pt* | 2.6 (1.2–4.2)* | 0.00 |
| *Longitudinality, per 10 pt* | 2.7 (1.1–4.4)* | 0.00 |
| *Coordination, per 10 pt* | 1.2 (0.1–2.4)* | 0.02 |
| *Comprehensiveness (service available), per 10 pt* | 1.9 (0.6–3.4)* | 0.007 |
| *Comprehensiveness(service provided), per 10 pt* | 1.5 (0.1–2.4)* | 0.004 |
| *Community Orientation, per 10 pt* | 2.4 (1.1–3.8)* | 0.00 |

Analysis of 194 patients among 29 facilities.
<sup>a</sup>Mixed-effects linear regression model adjusted for covariates listed in Table 4.
**\*statistically significant**

## Discussion

This study examined the association of quality of patient-centered care with QOL-HC and HR-Hope among patients receiving home medical care. Higher levels of patient centeredness, as measured using the JPCAT-SF, were associated with better QOL-HC and higher HR-Hope. In particular, the “first contact” and “longitudinality” domains of the JPCAT-SF were strongly associated with QOL-HC and HR-Hope.

Previous studies have also highlighted the importance of first contact and longitudinality in fostering hope through effective communication. For example, research involving psychologically ill patients receiving home medical care found that hope was associated with patients having sufficient time to communicate with their doctors and address their concerns.^9^ Additionally, studies involving terminally ill patients with malignancies have emphasized the role of communication in maintaining hope.^5^ However, the concepts and measurements of hope in these studies have’ not been structured or validated. Furthermore, the limitations of these studies include their specific focus on psychology and end-of-life patients with malignancies, which restrict the generalizability of their findings. Previous studies have shown that patient-centered care improves QOL in different populations, including outpatients with type 2 diabetes and nursing home residents.^3,4^

The results of the present study have several clinical implications. First, within the JPCAT domains, first contact and longitudinality were found to have stronger associations with QOL-HC and HR-Hope compared with other domains. These domains closely align with the communication pathway, particularly “access to care” and “enhancing therapeutic alliance.” Improving first contact by providing better access to care based on patient requests and understanding patients as individuals within the context of their life histories can significantly impact QOL and hope. Second, while previous studies have focused on developing additional programs, such as psychosocial supportive interventions, to improve patients’ hope, this study highlights the importance of enhancing patient centeredness in daily clinical practice. Improving daily clinical practice by prioritizing patient-centered conversations is more crucial than developing new programs. From this perspective, for example, a patient-centered conversation program developed for patients with chronic kidney disease to promote advanced care planning that promotes sharing patients’ values and preferences about treatment, their families, and everyday life among medical providers, patients, and their families could be applied to home medical care.^15^

This study had several strengths. First, using validated scales, we could quantify the associations between patient centeredness, QOL, and hope for the first time. Second, our findings are generalizable because of the multicenter nature of the study, which was conducted in both rural and urban areas of Japan. Third, by controlling for confounding variables, such as life expectancy and depressive symptoms, we could correctly estimate the associations of patient-centered care with QOL and hope.

Nonetheless, this study has several limitations. First, the possibility of reverse causation remains because of the study’s cross-sectional design. For example, because patients were highly hopeful about their health, their home physicians may have responded to this and provided them with good patient-centered care. Second, because this study excluded patients with severe dementia and those who were unable to respond, the findings may not be generalizable to these populations.

## Conclusion

In conclusion, our study revealed that better patient-centered care was associated with a higher QoL and hope among patients receiving home medical care. Further empirical research is warranted to determine whether efforts to holistically understand patients and promptly provide timely home medical care tailored to patients’ individual needs can improve patients’ overall well-being and hope.

## Data Availability

All data produced in the present work are contained in the manuscript.

