## Supplementary Files for "The impact of patient-centered care on quality of life and hope among patients receiving home medical care: The Zaitaku Evaluative Initiatives and Outcome Study"

**Supplementary infomation**

### **Supplementary Table S1. Items and responses for the JPCAT-SF**

Questionnaires in Japanese version are available from the following website (<https://bfffe681-45f7-48c3-aa93-2f67612434a5.filesusr.com/ugd/6c0e9c_7c80f3e3ce6e45eebb5a3aedfb9a2800.pdf>).

| **Instruction sentences** | **(English: “Check the box that best fits each question.”)** |
| --- | --- |
| Question 1 | (English: “When your Primary Care Practice is closed on Saturday and Sunday and you get sick, would someone from there see you the same day?”) |
| Response to Question 1 | (English: “Strongly agree/Somewhat agree/Not sure/Somewhat disagree/Strongly disagree ”) |
| Question 2 | (English: “When your Primary Care Practice is closed and you get sick during the night, would someone from there see you that night?”) |
| Response to Question 2 | (English: “Strongly agree/Somewhat agree/Not sure/Somewhat disagree/Strongly disagree ”) |
| Question 3 | (English: “Does your Primary Care Physician (PCP) know you very well as a person, rather than as someone with a medical problem?”) |
| Response to Question 3 | (English: “Strongly agree/Somewhat agree/Not sure/Somewhat disagree/Strongly disagree ”) |
| Question 4 | (English: “Does your PCP know what problems are most important to you?”) |
| Response to Question 4 | (English: “Strongly agree/Somewhat agree/Not sure/Somewhat disagree/Strongly disagree ”) |
| Question 5 | (English: “Have you ever had a visit to a specialist or special service of any kind?”) |
| Response to Question 5 | (English: “Yes/No or Not sure”) |
| Question 6 | (English: “Did your PCP suggest you go to the specialist or special service?”) |
| Response to Question 6 | (English: “Strongly agree/Somewhat agree/Not sure/Somewhat disagree/Strongly disagree ”) |
| Question 7 | (English: “Did your PCP discuss with you the different places you could have visited to get help with that problem?”) |
| Response to Question 7 | (English: “Strongly agree/Somewhat agree/Not sure/Somewhat disagree/Strongly disagree ”) |
| Question 8 | (English: “Please indicate whether it is available at your PCP’s office. Counselling related to abuse”) |
| Response to Question 8 | (English: “Strongly agree/Somewhat agree/Not sure/Somewhat disagree/Strongly disagree ”) |
| Question 9 | (English: “Please indicate whether it is available at your PCP’s office. Counselling related to personal preferences about end-of-life issues”) |
| Response to Question 9 | (English: “Strongly agree/Somewhat agree/Not sure/Somewhat disagree/Strongly disagree ”) |
| Question 10 | (English: “In visits to your PCP, are any of the following subjects discussed with you? Advice about over-the-counter medications.”) |
| Response to Question 10 | (English: “Strongly agree/Somewhat agree/Not sure/Somewhat disagree/Strongly disagree ”) |
| Question 11 | (English: “In visits to your PCP, are any of the following subjects discussed with you? Advice about medical information in the media: on TV, in the newspaper, etc.”) |
| Response to Question 11 | (English: “Strongly agree/Somewhat agree/Not sure/Somewhat disagree/Strongly disagree ”) |
| Question 12 | (English: “Does your PCP investigate whether the available health care is meeting the needs of the community?”) |
| Response to Question 12 | (English: “Strongly agree/Somewhat agree/Not sure/Somewhat disagree/Strongly disagree ”) |
| Question 13 | (English: “Does your PCP investigate the concerns people have about health problems in your community?”) |
| Response to Question 13 | (English: “Strongly agree/Somewhat agree/Not sure/Somewhat disagree/Strongly disagree ”) |

Each domain consists of the following items:

First contact domain - Questions 1 and 2

Longitudinality domain - Questions 3 and 4

Coordination domain - Questions 5, 6, and 7

Comprehensiveness (services available) domain - Questions 8 and 9

Comprehensiveness (services provided) domain - Questions 10 and 11

Community orientation domain - Questions 12 and 13

#### Scoring

For each item, participants were asked to respond on a 5-point Likert scale, ranging from *strongly disagree* to *strongly agree*. Each response was converted to an item score ranging from 0 to 4. The domain scores were calculated by multiplying the average of the item scores in the same domain by 25 (i.e., ranging from 0 to 100), with higher scores indicating better performance. In the coordination domain, which asks about experiences with referrals to a specialist, respondents who had never seen a specialist were given 50 points (the midpoint of all the possible scores). The total score was the average of the six domain scores and represented an overall measure of the patient experience of primary care.

**Supplementary Table S2. QOL for patients receiving home-based medical care. (QOL-HC).**

| Question 1 | (English: “Do you have peace of mind?”) |
| --- | --- |
| Question 2 | (English: “Do you feel satisﬁed with your life when you reﬂect on it?”) |
| Question 3 | (English: “Do you have someone that you spend time talking with?”) |
| Question 4 | (English: “Are you satisﬁed with the home care service system?”) |
| Response options for Question | (English: Never agree (0)/Neither agree nor disagree (1)/always agree (2)) |

The original English version is provided for each item and response.

**Supplementary Table S3. Japanese version of the health-related hope scale (HR-Hope).**

| In the past 30 days, how much difficulty did you have in: | |
| --- | --- |
| Question 1 | (English: “I think I’ll still be able to continue doing enjoyable things in the future.”) |
| Question 2 | (English: “I will probably be able to discover some meaning to my life.”) |
| Question 3 | (English: “I will probably be able to work, to the best of my ability, to find a sense of purpose in life.”) |
| Question 4 | (English: “I will probably be able to live each day to its fullest.”) |
| Question 5 | (English: “I feel that I can continue to experience a sense of fulfillment in my daily life.”) |
| Question 6 | (English: “Even if I should feel down due to my illness, I could probably turn my feelings around.”) |
| Question 7 | (English: “I feel I can adjust my health goals in a way that is consistent with my actual disease condition.”) |
| Question 8 | (English: “I can probably alter my goals depending on changes in my illness or symptoms”) |
| Question 9 | (English: “Even if my health condition keeps me from achieving my present goals, I will probably be able to find a new goal.”) |
| Question 10 | (English: “I can probably develop a personal lifestyle strategy for dealing with my disease condition.”) |
| Question 11 | (English: “I will probably be able to find a way to keep my illness from worsening.”) |
| Question 12 | (English: “I will probably be able to continue performing my role in society”) |
| Question 13 | (English: “My disease experience will probably encourage those around me to be mindful of their own health.”) |
| Question 14 | (English: “I feel I can deepen my relationships with my friends.”) |
| Question 15 | (English: “Those around me will probably go along with any changes in my mood.”) |
| Question 16 | (English: “Those around me will probably continue to treat me the same way they always have”) |
| Question 17* | (English: “I’ll probably be able to continue my usual role in support of my family.”) |
| Question 18* | (English: “I feel that I’ll continue to have a good relationship with my family.”) |
| Response options  for Questions | (English: I don’t feel that way at all (0)/ I feel that way a little (1)/ I feel that way somewhat (2)/ I feel that way strongly (3) |

*Questions for people with families

When using this instrument, please refer to the following reference.

**Reference**

Fukuhara S, Kurita N, Wakita T, Green J, Shibagaki Y.

A scale for measuring health-related hope: its development and psychometric testing.

Annals of Clinical Epidemiology 2019;1(3):102–119

**Supplementary Text S1. Description of the psychometric properties of the JPCAT-SF and the concepts of its domains**

The JPCAT-SF is a short version of the original 29-item JPCAT,^1^ which was itself an adaptation to the Japanese culture of the Primary Care Assessment Tool (PCAT) designed to measure the experience of adult patients in primary care.^2^ In an outpatient setting, the JPCAT-SF has been shown to have good internal-consistency reliability (Cronbach's α = 0.77 for the total score, Cronbach's α > 0.76 for each domain score) and excellent criterion validity (Pearson correlation coefficient with the original 29-item JPCAT and the overall rating for usual care facilities: 0.94 and 0.43, respectively).^3^

First contact

Care is first sought from a primary care provider when a new health or medical need arises. The service should also be accessible and usable by the population as a new need or problem arises.^4^ First contact is closely related to “access to care,” a domain of patient-centred care characterized by the timely availability of care that is tailored to the patient.^5^ The JPCAT-SF mainly measures patient experience related to off-hours care in primary care.^4^

Longitudinality

It refers to the longitudinal use of usual sources of care, regardless of illness or injury.^4^ Longitudinality is supported by one of the principles for patient-centeredness, namely the consideration of the “patient as a unique person,” i.e., the primary care physician's recognition of the patient's uniqueness (individual needs, preferences, values, beliefs, concerns, etc.).^5^ The JPCAT-SF mainly measures whether a patient feels that their primary care physician recognizes them as a whole person.^4^

Coordination

The essence of coordination is the availability of information about past and existing problems and services and the recognition of that information in relation to a current care need.^4^ It relates to “coordination and continuity of care,” which is an enabler of patient-centred care, i.e., facilitation of care that is well-coordinated and continuous.^5^ The JPCAT-SF mainly measures patient experience regarding past referrals to a specialist.^4^

Comprehensiveness (services available)

It refers to the availability of a wide range of services by a primary care provider and their appropriateness for a spectrum of needs for all but the most uncommon problems.^4^

Under "services available," the JPCAT-SF mainly measures whether a patient feels they can receive care for mental health, dementia, and advanced care planning, if necessary.^4^

Comprehensiveness (services provided)

It includes appropriate advice on daily lifestyle habits, self-medication, and health literacy.^4,6^ It is underpinned by patient empowerment, an activity of patient-centeredness, in which a primary care physician recognizes and actively supports a patient's ability and responsibility to self-manage their illness.^5^ The JPCAT-SF mainly measures patient experience in terms of whether they received such appropriate advice.

Community orientation

It refers to care that is delivered in the context of the community ^4^ and is considered as a derivative domain of principles of primary care.^1^

The JPCAT-SF mainly measures patient experience regarding home visits and whether a patient feels that their primary care physician is interested not only in their individual health problem but also in problems in the community.^4^

### **Supplementary Text S2. Full details of the other members of the ZEVIOUS Group**

Shinsuke Muto^1^; Tatsunobu Natsubori^1,2^; Michiko Hinata^1,3^; Wataru Nakagawa^1^; Akihiko Yonenaga^1^; Lina Inagaki^1^; Shioto Itakura^1^; Nobuhiro Ikeda^1^; Tomoka Nakamura^1^; Naoya Miyashita^1^; Takuya Furugen^1^; Takafumi Abo^4,5^; Sadayuki Okudaira^4,6^; Kazuhiko Takuma^4,7^; Chihiro Tsuchiya^,8^; Masahiro Deguchi^4,9^; Takashi Fujii^4,10^; Yoshitaka Harada^4,11^; Seiji Matsuo^4,12^; Motomichi Nakagawa^4,13^; Ken Tanigawa^4,14^; Yoshio Ochi^4,15^; Sadanobu Ogasawara^4,16^; Kazuhiko Hoshino^4,17^; Momoko Aruga^18^; Yoshinori Nakamura^18^; Nobuhiro Sawa^19^; Yosuke Akashi^19^; Nobuyuki Miyagi^20^; Toyohiro Terasaki^21^; Kunihiro Kinoshita^22^; Masaji Kikukawa^23^; Hisakazu Kato^24^; Masayuki Amano^25^; Kentaro Asakura^26^; and Naoto Fukui^27^.

^1^You Home Clinic, Bunkyo City, Japan

^2^You Home Clinic Azumabashi, Sumida City, Japan

^3^You Home Clinic Azabudai, Minato City, Japan

^4^Dr. Net Nagasaki, Nagasaki City, Japan

^5^Abo Gastrointestinal Surgical Clinic, Nagasaki City, Japan

^6^Okudaira Geka, Nagasaki City, Japan

^7^Takuma Clinic, Nagasaki City, Japan

^8^Chihiro Naika Clinic, Nagasaki City, Japan

^9^Deguchi Surgery Clinic, Nagasaki City, Japan

^10^Fujii Surgical Clinic, Nagasaki City, Japan

^11^Harada Internal Medicine Clinic, Nagasaki City, Japan

^12^Nagasaki Takara Home Medical Care Clinic, Nagasaki City, Japan

^13^Nakagawa Surgical Clinic, Nagasaki City, Japan

^14^Tanigawa Clinic, Nagasaki City, Japan

^15^Ochi Clinic, Nagasaki City, Japan

^16^Nagasaki Memorial Hospital, Nagasaki City, Japan

^17^Hoshino Internal and Respiratory Medical Clinic, Nagasaki City, Japan

^18^Medical home care center, Tenri Hospital Shirakawa Branch, Tenri City, Japan

^19^Minami Nara General Medical Center, Oyodo, Japan

^20^Miyagi Clinic, Tenri City, Japan

^21^Terasaki Clinic, Nara City, Japan

^22^Kinoshita Clinic, Sakurai City, Japan

^23^Kikukawa Internal Medicine Clinic, Sakurai City, Japan

^24^Kato Clinic, Uda City, Japan

^25^Nosegawa Village National Health Insurance Clinic, Nosegawa, Japan

^26^Daifuku Clinic, Sakurai City, Japan

^27^Fukui Clinic, Uda City, Japan
